# Normative Values of Echocardiographic Chamber Size and Function in Older Healthy Adults: The Multi-Ethnic Study of Atherosclerosis

**DOI:** 10.1101/2023.12.05.23299572

**Authors:** Monica Mukherjee, Jordan B. Strom, Jonathan Afilalo, Mo Hu, Lauren Beussink-Nelson, Jiwon Kim, Karima Addetia, Alain Bertoni, John Gottdiener, Erin D. Michos, Julius M. Gardin, Sanjiv J. Shah, Benjamin H. Freed

**Author notes:** Signifies co-first authors (Monica Mukherjee, Jordan B. Strom). **Address correspondence to:** *+signifies co-corresponding authors*. Monica Mukherjee, MD, MPH, Johns Hopkins University School of Medicine, 301 Mason Lord Drive, Suite 2400, Baltimore, MD 21224. Benjamin H. Freed, MD, Northwestern Feinberg School of Medicine, Galter Pavilion, 675 N St Clair St #19-100, Chicago, IL 60611.

## Abstract

**Background:** Echocardiographic (2DE) thresholds indicating disease or impaired functional status compared to normal physiologic aging in individuals ≥ 65 years are not clearly defined. In the present study, we sought to establish standard values for 2DE parameters related to chamber size and function in older adults without cardiopulmonary or cardiometabolic conditions.

**Methods:** In this cross-sectional study of 3032 individuals who underwent 2DE at Exam 6 in the Multi-Ethnic Study of Atherosclerosis (MESA), 608 participants fulfilled our inclusion criteria, with normative values defined as the mean value ± 1.96 standard deviations and compared across sex and race/ethnicity. Functional status measures included NT-proBNP, 6-minute walk distance [6MWD], and Kansas City Cardiomyopathy Questionnaire [KCCQ]. Prognostic performance using MESA cutoffs was compared to established guideline cutoffs using time-to-event analysis.

**Results:** Participants meeting our inclusion criteria (69.5 ± 7.0 years, 46.2% male, 47.5% White) had lower NT-proBNP, higher 6MWD, and higher (better) KCCQ summary values. Women had significantly smaller chamber sizes and better biventricular systolic function. White participants had the largest chamber dimensions, while Chinese participants had the smallest, even after adjustment for body size. Current guidelines identified 81.6% of healthy older adults in MESA as having cardiac abnormalities.

**Conclusions:** Among a large, diverse group of healthy older adults, we found significant differences in cardiac structure and function across sexes and races/ethnicities, which may signal sex-specific cardiac remodeling with advancing age. It is crucial for existing guidelines to consider the observed and clinically significant differences in cardiac structure and function associated with healthy aging. Our study highlights that existing guidelines, which grade abnormalities in echocardiographic cardiac chamber size and function based on younger individuals, may not adequately address the anticipated changes associated with normal aging.

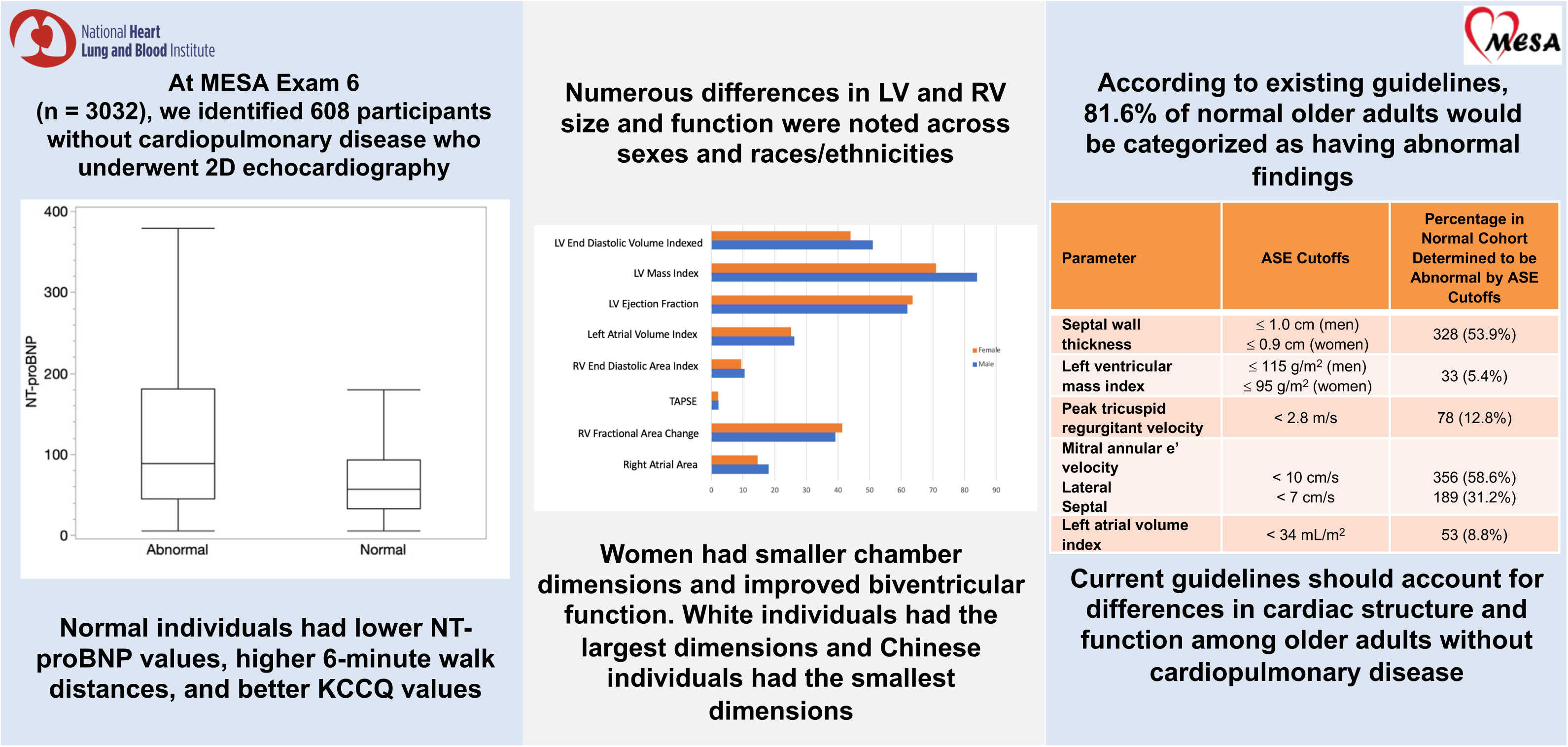

## INTRODUCTION

Two-dimensional echocardiography (2DE) has a well-established role in the comprehensive assessment of cardiac size and function with standardized methodologies to allow for epidemiologic comparisons within and between individuals. Moreover, the judgment of whether an anatomic structure is within normal limits on 2DE has a significant public health impact on treatment assignments, downstream utilization, and healthcare expenditures. While several prior studies have aimed to define normative values based on race and ethnicity ^1–8^, there is limited understanding regarding alterations in these normative values due to physiologic aging, a process associated with progressive alterations in cardiac structure and function ^9^.

Physiologic aging is associated with progressive changes in cardiac structure and function due to vascular stiffening ^10^ and sex-specific dimorphisms ^9, 11^, and mechanistically related to increased collagen deposition and fibrosis. Early studies established a pattern of increasing left ventricular (LV) stiffness, reduced diastolic compliance, and altered LV filling characteristics, with increased atrial contribution with physiologic aging ^12, 13^. Despite the known effects of age on cardiac remodeling, guideline-recommended echocardiographic normative values ^14, 15^ have been derived from epidemiologic studies, predominantly enrolling young individuals without cardiovascular disease (CVD). Study cohorts are also limited by the small proportion of older individuals, limited ethnic and racial diversity, and lack of the correlation of normal physiologic aging on these parameters and the association with incident heart failure (HF) and functional measures such as six-minute walk distance (6MWD).

The Multi-Ethnic Study of Atherosclerosis (MESA) Heart Failure is an ancillary study of MESA participants who underwent state-of-the-art echocardiography at Exam 6. In this analysis, we evaluated a large, diverse, well-characterized, community-based representative cohort of older adults without cardiopulmonary or cardiometabolic disease to 1) determine if cardiac chamber size and function differ by age, sex, race/ethnicity; 2) establish normative values and grades of severity; 3) evaluate whether individuals without prevalent cardiopulmonary or cardiometabolic disease (i.e., normative aging cohort) have essential differences in functional measures such as N-terminal pro-B-type natriuretic peptide (NT-proBNP), 6MWD, and Kansas City Cardiomyopathy Questionnaire (KCCQ) score compared to those not included in the normative aging cohort ^16^; and 4) determine whether the observed risk of adverse outcomes is similar when individuals are categorized according to guideline vs. MESA cutoffs. We hypothesize that there are both sex-and race-specific differences with cardiac remodeling that occur with age, which may have significant implications for echocardiographic grading of abnormality in older adults.

## METHODS

### Transparency and Openness Policy

Data from MESA are available through the National Heart, Lung, Blood Institute (NHLBI) Biologic Specimen and Data Repository Coordinating Center. Requests for access to the data are made through the website: https://biolincc.nhlbi.nih.gov/studies/mesa.

### Study Population

MESA is a prospective multicenter study of 6814 participants aged 45-84 years old at baseline across six communities within the United States encompassing varying demographic groups^17^. At the baseline examination (2000-2002), all MESA participants were free of clinical CVD, including HF, and prospectively followed for incident CVD events through 2019 through annual telephone calls. The MESA protocol, information about the source populations from which recruitment occurred, detailed exclusion criteria, investigator contact details, and other information are available at www.mesa-nhlbi.org. Each field side obtained an institutional review board agreement, and all participants provided written consent.

In an ancillary study designed to investigate early signs of HF, conducted at examination 6, a comprehensive 2DE was performed in 3032 participants. We defined a normative aging cohort of these individuals by excluding participants with known cardiopulmonary or cardiometabolic disease, characterized as prevalent or incident adjudicated HF or coronary heart disease events at or between Exam 1 and Exam 6, shown in **Figure 1** ^18, 19^. Our exclusion criteria included: history of myocardial infarction (n=110), history of clinical HF (n=68), biplane LV ejection fraction (LVEF) <50% as measured on 2DE at Exam 6 (n= 40), use of antihypertensive medications (n=1680), current smoking (n=58), history of diabetes (n=28), hemoglobin A1c ≥ 7 (n=9), history of non-sinus rhythm (n=76), history of chronic kidney disease (CKD, n=8), estimated glomerular filtration rate (eGFR), calculated by the chronic kidney disease epidemiology collaboration formula,^20^ ≤ 60 mL/min/1.73m^2^ (n=71), history of chronic obstructive pulmonary disease (n=7), history of asthma (n=27), history of chronic respiratory disease (n=7), history of hypertension (HTN, n = 211), systolic blood pressure (SBP) ≥ 140 mmHg (n = 0 after applying other exclusions), or aortic valve area ≤ 1.5 cm2 (n=24).

**Figure 1.**
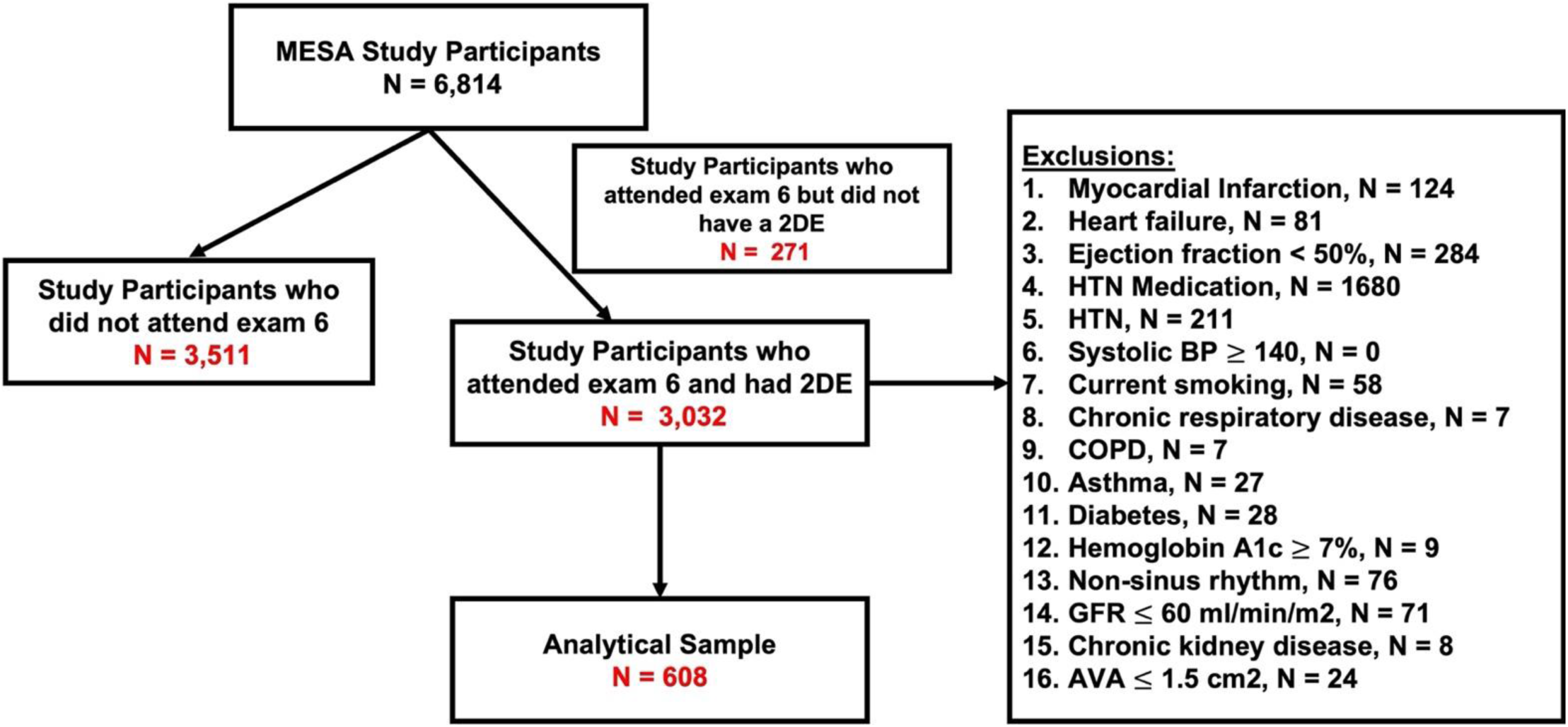
Flowchart of Study Participants and Inclusion and Exclusion Criteria.

### Functional Status Measures

Functional status markers of cardiovascular function included NT-proBNP, 6MWD, and KCCQ metrics. NT-proBNP was measured by an Elecsys immunoassay (Roche Diagnostics, Indianapolis, IN)^21^ in a subset of MESA participants (n = 1000). Ambulatory MESA Exam 6 participants were invited to undergo 6MWD testing (n = 2539) using a standardized protocol^22^, and encouraged to walk as quickly as possible on a measured track for a timed 6 minutes.

6MWD was defined as the total distance walked (in meters) in 6 minutes or until the participant requested to stop. The validated KCCQ survey ^23^ was administered to 3097 individuals at Exam 6 to assess quality of life.

### Echocardiography

2DE were performed using dedicated Vivid T8 ultrasound systems (GE Healthcare, Waukesha, WI) during MESA Examination 6 (2016–2018) from each study site and analyzed at the Northwestern Echocardiography Core Laboratory using the EchoPAC platform (GE Healthcare, Chicago, IL). Linear and volumetric imaging of cardiac chambers were obtained using 2DE-directed methods, and color, pulsed-and continuous-wave Doppler measurements were used to assess valvular function, following American Society of Echocardiography (ASE) guidelines ^24, 25^. LV diastolic parameters included early (E) and late (A) diastolic inflow velocities, deceleration time, and tissue Doppler imaging (TDI) medial and lateral e’ velocities ^14, 15^. Tricuspid annular plane systolic excursion (TAPSE), fractional area change (FAC), and TDI peak systolic (S’) velocity metrics were used to assess right ventricular (RV) systolic function. ^26^. FAC was calculated using RV end-diastolic and end-systolic areas ^26^. Applying the modified Bernoulli equation, RV systolic pressure (RVSP) was estimated utilizing the peak tricuspid regurgitant (TR) velocity and adding estimated RA pressure based on inferior vena cava dimension (IVC) and collapsibility to calculate pulmonary arterial systolic pressure (PASP) ^27, 28^.

### Covariate Assessment

Covariates were included from the MESA 6 visit, during which study participants underwent standardized questionnaires, physical examinations, and comprehensive laboratory testing. In the present analysis, we considered demographic factors (age, sex, race/ethnicity, MESA field site, behavioral factors (smoking status and physical activity), anthropometric factors (height, weight, and body surface area ^29^, body mass index [BMI]^30^), cardiovascular risk factors such as abnormal SBP, abnormal kidney function, and presence of diabetes. Height and weight were measured at the time of 2DE. The presence of diabetes was determined if the fasting blood glucose level was ≥126 mg/dL, a self-reported diagnosis of diabetes, or current use of diabetes medications. We also included the use of antihypertensive medications. Laboratory studies used for this analysis included the eGFR, calculated by the chronic kidney disease epidemiology collaboration formula^20^.

### Outcome Measures

Prognostic performance was determined by comparing the outcomes of participants with abnormal values by MESA-defined cutoffs to those of ASE-guideline-defined cutoffs. Individuals with values exceeding the cutoff value (**Supplemental Table 1**) for any of the 12 echocardiographic parameters (left atrial end-systolic volume index, LV diastolic volume index, LV ejection fraction, LV mass index, septal wall thickness, TDI medial and lateral e’ velocities, average E/e’, RV end-diastolic area index, TAPSE, FAC, and peak TR velocity) were considered as having an abnormality in cardiac structure or function for comparison.

Participants were followed from visit 1 through December 31, 2019, for any of the following events: myocardial infarction (MI), percutaneous coronary intervention (PCI), coronary artery bypass grafting (CABG), revascularization (the composite of PCI or CABG), cerebral vascular accident (CVA), transient ischemic attack (TIA), heart failure, death, and a composite of MI, revascularization, CVA, TIA, or heart failure. A separate clinical events committee performed detailed event adjudication. Definitions for these endpoints and detailed information regarding event ascertainment are available at www.mesa-nhlbi.org.

### Statistical Analysis

Variables were described using means, standard deviations (SD), counts, and proportions as appropriate, employing a cross-sectional study design. The normality of measured variables was verified using Q-Q plots, histograms, and evaluation of skewness/kurtosis measures. Characteristics of participants meeting inclusion criteria (i.e., the normative aging cohort) were compared to those not meeting inclusion criteria (the non-normative aging cohort) using t-tests or Fisher’s exact tests. Box and whisker plots were used to display the distribution of NTproBNP, 6MWD, and KCCQ values between the subgroups, and differences in the values between these groups were evaluated using t-tests. In the combined normative and non-normative aging cohorts, nested multivariable linear regression models were used to assess the relationship between normal cohort inclusion status and NT-proBNP, 6MWD, and KCCQ, first adjusting for age, sex, race/ethnicity, and site of enrollment (Model 1) and subsequently adjusting for these variables plus body surface area and waist circumference (Model 2).

Subsequently, echocardiographic measures were stratified by sex and compared across sex categories using t-tests, and additionally stratified by race/ethnicity and compared across race/ethnicity categories using analysis of variance (ANOVA). In all stratified analyses, normal ranges were defined as upper and lower limits using 1.96 SDs from the mean value in the normative aging population (N = 608). The degree of abnormality was determined using the following grading system: mild [mean ± 2-3 SDs], moderate [mean ± 3-4 SDs, and severe mean ± 4 or more SDs]. According to current ASE guidelines, the proportion of individuals in the normative aging cohort classified as abnormal was determined for five parameters (septal wall thickness, LV mass index, peak TR velocity, mitral annular e’ velocity, and LA volume index). In a sensitivity analysis, we excluded those with septal wall thickness ≥ 1.5 cm to assess how this influenced results, as this may reflect the measurement of basal septal thickening in older adults.

We then calculated survival times using the Exam 6 date as the start date and defined four outcomes to be analyzed: all incident CVD using established MESA definitions, incident HF, death, and incident HF or death. If a participant had the outcome before Exam 6, they were excluded from the analysis for that particular outcome. We further compared the receiver-operating area under the curve (AUC), sensitivity, specificity, positive predictive value (PPV), and negative predictive value (NPV) between ASE and MESA cut-offs. AUC p-values for testing were performed if there was a significant difference in the Brier scores^31^, a measure of the accuracy of probabilistic predictions, between the null and Cox models. We performed McNemar’s Test to generate p-values from the calculation of the confusion matrix, testing that the proportions of false positives and false negatives are equal.

The reproducibility of measurements was evaluated using intraclass correlation coefficients (ICCs). Intra-observer and inter-observer variability for all 2DE measurements ranged from 0.80-0.99 for all measurements (**Supplemental Review File**). We used STATA version 15.0 (StataCorp LP, College Station, TX), R version 3.5.1 (R Foundation for Statistical Computing) and JMP v15.0 (SAS Institute, Cary, NC) for statistical analysis. Statistical significance was defined by a two-sided p-value<0.05.

## RESULTS

Of 3032 participants who underwent 2DE at MESA Exam 6, 608 participants met inclusion criteria for the normative aging cohort (**Figure 1**) and had a mean age of 69.5 ± 7.0 years, and 46.2% were male, 47.5% were White, 17.4% Chinese, 14.5% Black, and 20.6% Hispanic race/ethnicity (**Table 1**). The mean BMI was 26.7 ± 5.0 kg/m^2^. The mean blood pressure was lower in women, p < 0.05, and non-included individuals (i.e., those in the non-normative aging cohort) were younger, more frequently White, or Chinese adults, had lower blood pressures, lower hemoglobin A1c values, higher eGFR values, performed greater amounts of moderate to vigorous exercise, were less likely to smoke tobacco, and were less frequently treated with cardiovascular medications (all p<0.001). Additionally, these participants had lower values of NT-proBNP and better exercise capacity and health status, as suggested by higher 6MWD and KCCQ scores, respectively, as shown in **Figure 2**. These group differences persisted despite adjustments for age, sex, race/ethnicity, enrollment site, body surface area, and waist circumference, **Supplemental Table 2**.

**Figure 2.**
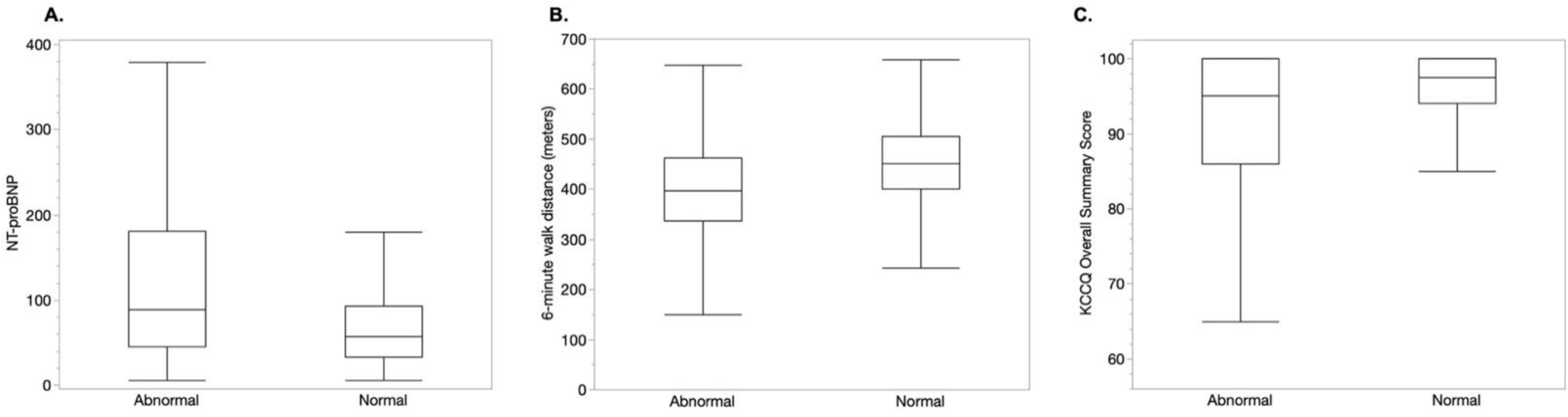
Graphical Findings of Normative Values Stratified by Sex, Race/Ethnicity, and Comparison of Cut-offs to Established Guidelines. Graphical representations are shown for **A.** normative values stratified by sex, **B.** normative values stratified by race and ethnicity, and **C.** cut-offs for normality comparing MESA normative values cohort and ASE guidelines.

**Table 1.** Clinical Characteristics of MESA Exam 6 Participants. Clinical characteristics of MESA study participants who underwent 2DE at Exam 6 were stratified by whether or not they met inclusion criteria for the normal cohort (n = 608) or not (n = 2424). P-values represent comparison between the normal vs. abnormal cohorts. Results are displayed as percentages or means ± standard deviations. *None of the MESA participants included in the sample meeting our inclusion criteria had atrial fibrillation at the time of 2DE. Two patients were taking Warfarin for a remote history of AF, however, were in normal sinus rhythm at the time of 2DE. Three patients were taking Warfarin for non-cardiac indications (presumably history of deep vein thrombosis and/or thromboembolism).

| Participant Characteristics | Number of Observations | Abnormal Cohort<br>n=2424 | Normal Cohort<br>n=608 | p-value |
| --- | --- | --- | --- | --- |
| Male sex | 3032 | 47.2% | 46.2% | 0.69 |
| Age at Exam 6 | 3032 |  |  | < 0.001 |
| 55-64 |  | 11.5% | 28.0% |  |
| 65-74 |  | 37.8% | 49.2% |  |
| 75-84 |  | 34.7% | 19.2% |  |
| 85 or older |  | 15.9% | 3.6% |  |
| Race/Ethnicity | 3032 |  |  | <0.001 |
| White |  | 37.9% | 47.5% |  |
| Chinese |  | 12.4% | 17.4% |  |
| Black |  | 27.8% | 14.5% |  |
| Hispanic |  | 21.9% | 20.6% |  |
| Height, cm | 3031 | 164.5 ± 10.1 | 165.9 ± 9.2 | 0.003 |
| Weight, lbs | 3031 | 173.2 ± 40.0 | 162.4 ± 35.5 | <0.001 |
| Waist Circumference, cm | 3009 | 100.7 ± 14.4 | 94.3 ± 12.9 | <0.001 |
| Body Mass Index, kg/m<sup>2</sup> | 3029 | 28.9 ± 5.8 | 26.7 ± 5.0 | <0.001 |
| Systolic Blood Pressure, mmHg | 3027 | 130.6 ± 21.3 | 115.3 ± 13.1 | <0.001 |
| Diastolic Blood Pressure, mmHg | 3027 | 69.1 ± 10.4 | 66.8 ± 8.4 | <0.001 |
| Heart rate, bpm | 3032 | 64.6 ± 11.2 | 64.2 ± 9.1 | 0.41 |
| Total Cholesterol, mg/dl | 2961 | 200.1 ± 38.1 | 181.7 ± 41.2 | <0.001 |
| Serum Hemoglobin A1c, % | 1797 | 6.0 ± 1.0 | 5.6 ± 0.4 | <0.001 |
| eGFR (CKD-EPI Equation) | 2961 | 71.8 ± 19.0 | 82.9 ± 11.5 | <0.001 |
| Presently Drink Alcohol | 3024 | 40.5% | 51.6% | <0.001 |
| Cigarette Smoking Status | 3024 |  |  | <0.001 |
| Never |  | 44.9% | 53.4% |  |
| Former |  | 48.1% | 46.6% |  |
| Current |  | 7.0% | 0.0% |  |
| Moderate to vigorous exercise | 3020 | 23.1% | 33.1% | <0.001 |
| Aspirin | 3025 | 46.3% | 25.2% | <0.001 |
| Insulin | 3019 | 4.8% | 0.0% | <0.001 |
| Inhaled steroids | 3019 | 0.3% | 0.2% | 0.80 |
| Warfarin Use* | 3019 | 2.8% | 0.8% | 0.008 |
| Any Lipid Lowering Medication | 3019 | 52.2% | 28.5% | <0.001 |
| Any Diuretic | 3019 | 22.4% | 0.0% | <0.001 |
| Any Hypertension Medication | 3019 | 76.7% | 0.0% | <0.001 |

Normative echocardiographic measures stratified by sex are shown in **Table 2**. Biplane LV chamber volumes were smaller in women, even when indexed for BSA. While LA volume differed between sexes (p<0.001), this difference was no longer significant when indexed for BSA. Female participants had smaller LV chamber wall thickness, mass, and LVOT diameters. Stroke volume and cardiac output (and respective indexed measures) were also smaller in female participants; however, LVEF was higher (p<0.001). While the mitral E/A ratio did not differ across sexes, both lateral and septal E/e’ ratios were higher in women (p<0.05 for both). Right heart parameters were also smaller in women, regardless of indexation for BSA. Female participants had higher FAC (p<0.001) despite similar TAPSE and RV TDI S’ velocity values. Hemodynamic measures did not differ between the sexes.

**Table 2.** Echocardiographic Characteristics of MESA Exam 6 Participants. Displayed are measured echocardiographic parameters in MESA Exam 6 amongst the normal cohort (n = 608) stratified by self-reported sex. *Represents a single cutoff for moderate to severe abnormalities in this parameter. P-values for t-tests for the comparison of mean parameter values across sexes are provided.

| Echocardiographic Parameters | MALE |  |  |  | FEMALE |  |  |  | p-value |
| --- | --- | --- | --- | --- | --- | --- | --- | --- | --- |
|  | Normal | Mildly Abnormal | Moderately Abnormal | Severely Abnormal | Normal | Mildly Abnormal | Moderately Abnormal | Severely Abnormal |  |
| Left Ventricular Structural and Functional Variables |  |  |  |  |  |  |  |  |  |
| Biplane LA end-systolic volume, ml | 51.0<br>[23.7-78.3] | 78.4-91.9 | 92.0-105.6 | >105.6 | 44.0<br>[21.3-66.6] | 66.7-78.0 | 78.1-89.3 | >89.3 | <0.001 |
| Biplane LA end-systolic volume/BSA, ml/m2 | 26.2<br>[13.7-38.8] | 38.9-45.2 | 45.3-51.7 | >51.7 | 25.2<br>[14.1-36.2] | 36.3-41.9 | 42.0-47.4 | >47.4 | 0.05 |
| Biplane LV end-diastolic volume, ml | 86.8<br>[50.3-123.3] | 123.4 -141.5 | 141.6 -159.7 | >159.7 | 66.6<br>[39.9-93.1] | 93.2-106.6 | 106.7 -119.9 | >119.9 | <0.001 |
| Biplane LV end-diastolic volume index, ml/m2 | 44.6<br>[28.7-60.4] | 60.5 - 68.3 | 68.4 - 76.3 | >76.3 | 38.3<br>[25.7 - 50.8] | 50.9 - 57.1 | 57.2 - 63.4 | >63.4 | <0.001 |
| Biplane LV ejection fraction, % | 61.9<br>[53.9-69.9] | 49.9-53.8 | 45.9-49.8 | <45.9 | 63.6<br>[56.1-71.0] | 52.3-56.0 | 48.6-52.2 | <48.6 | <0.001 |
| Left ventricular end diastolic dimension, cm | 4.4<br>[3.4-5.4] | 5.5-5.9 | 6.0-6.4 | >6.4 | 4.1<br>[3.1-5.1] | 5.2-5.6 | 5.7-6.1 | >6.1 | <0.001 |
| Left ventricular end systolic dimension, cm | 3.0<br>[2.2-3.8] | 3.9-4.2 | 4.3-4.6 | > 4.6 | 2.7<br>[2.1-3.3] | 3.4-3.6 | 3.7-3.9 | >3.9 | 0.009 |
| Left ventricular mass, gm | 163.8<br>[83.1-244.5] | 244.6-284.8 | 284.9-325.2 | > 325.2 | 124.1<br>[60.3-187.9] | 188.0-219.8 | 219.9-251.7 | >251.7 | <0.001 |
| Left ventricular mass index, gm/m2 | 83.9<br>[48.7 - 119.1] | 119.2-136.7 | 136.8-154.3 | > 154.3 | 71.0<br>[41.4 - 100.5] | 100.6 -115.3 | 115.4 -130.1 | >130.1 | <0.001 |
| Left ventricular outflow tract diameter, cm | 2.21<br>[1.89-2.53] | 2.54-2.69 | 2.70-2.85 | > 2.85 | 1.97<br>[1.68-2.27] | 2.28-2.42 | 2.43-2.56 | >2.56 | <0.001 |
| Septal wall thickness, cm | 1.17<br>[0.75-1.60] | 1.61-1.81 | 1.82-2.03 | > 2.03 | 1.02<br>[0.59-1.46] | 1.47-1.67 | 1.68-1.89 | >1.89 | <0.001 |
| Posterior wall thickness, cm | 0.95<br>[0.69-1.21] | 1.22-1.34 | 1.35-1.47 | > 1.47 | 0.86<br>[0.63-1.08] | 1.09-1.20 | 1.21-1.31 | >1.31 | <0.001 |
| Fractional shortening, % | 32.9<br>[25.6-40.2] | 21.9-25.5 | 18.3-21.8 | < 18.3 | 33.9<br>[26.9-40.9] | 23.4-26.8 | 19.9-23.3 | <19.9 | 0.001 |
| Stroke work, g*m | 90.2<br>[40.9-139.5] | 16.2-40.8 | < 16.2* |  | 70.0<br>[30.3-109.1] | 10.6-30.2 | < 10.6* |  | <0.001 |
| LVOT VTI, cm | 22.1<br>[14.1-30.1] | 10.1-14.0 | 6.1-10.0 | < 6.1 | 23.2<br>[14.5-31.9] | 10.1-14.4 | 5.7-10.0 | < 5.7 | 0.001 |
| Stroke volume, ml | 84.9<br>[47.2-122.5] | 28.4-47.1 | 9.6-28.3 | < 9.6 | 70.8<br>[39.1- 102.6] | 23.2-39.0 | 7.3-23.1 | < 7.3 | <0.001 |
| Stroke volume index, ml/m2 | 43.7<br>[25.6-61.8] | 16.6-25.5 | 7.6-16.5 | < 7.6 | 40.8<br>[24.1-57.4] | 15.8-24.0 | 7.5-15.7 | < 7.5 | 0.002 |
| Cardiac output, L/min | 5.23<br>[2.58-7.87] | 1.26-2.57 | < 1.26* |  | 4.49<br>[2.24- 6.74] | 1.11-2.23 | < 1.11* |  | <0.001 |
| Cardiac index, L/min/m2 | 2.69<br>[1.42-3.96] | 0.79-1.41 | 0.16-0.78 | < 0.16 | 2.59<br>[1.34-3.83] | 0.72-1.33 | 0.09-0.71 | < 0.09 | <0.001 |
| Left ventricular ejection time, ms | 310.5<br>[262.0-359.1] | 237.7-261.9 | 213.4-237.6 | <213.4 | 319.6<br>[275.0-364.2] | 252.7 -274.9 | 230.4 -252.6 | < 230.4 | <0.001 |
| End-systolic elastance, mmHg/ml | 2.34<br>[1.19-3.49] | 0.62-1.18 | 0.04-0.61 | < 0.04 | 2.78<br>[1.36-4.21] | 0.64-1.35 | -0.07-0.63 | < -0.07 | 0.04 |
| Transmitral E-wave velocity, m/s | 69.2 | 98.9-113.5 | 113.6-128.3 | > 128.3 | 76.4 | 107.5-122.9 | 123.0-138.4 | > 138.4 | <0.001 |
|  | [39.7-98.8] |  |  |  | [45.3-107.4] |  |  |  |  |
| Transmitral A-wave velocity, m/s | 74.7<br>[38.7-110.7] | 110.8-128.7 | 128.8-146.7 | > 146.7 | 80.6<br>[45.3-115.9] | 116.0-133.6 | 133.7-151.3 | > 151.3 | <0.001 |
| E/A ratio | 0.97<br>[0.42-1.51] | 1.52-1.78 | 1.79-2.06 | > 2.06 | 0.99<br>[0.43-1.54] | 1.55-1.82 | 1.83-2.09 | > 2.09 | 0.39 |
| Mitral E-wave deceleration time, msec | 202.5<br>[118-287] | 288-329 | 330-371 | > 371 | 193.6<br>[117 – 271] | 272-309 | 310-347 | > 347 | 0.007 |
| Tissue Doppler Septal e' velocity, cm/s | 7.9<br>[4.2-11.6] | 2.3-4.1 | 0.5-2.2 | < 0.5 | 8.3<br>[4.2-12.5] | 2.1-4.1 | < 2.1* |  | 0.012 |
| Tissue Doppler Lateral e' velocity, cm/s | 9.7<br>[5.1-14.4] | 2.7-5.0 | 0.4-2.6 | < 0.4 | 9.6<br>[5.4-13.9] | 3.2-5.3 | 1.1-3.1 | < 1.1 | 0.554 |
| Tissue Doppler Septal a' velocity, cm/s | 11.5<br>[7.9-15.1] | 6.1-7.8 | 4.3 - 6.0 | < 4.3 | 10.6<br>[6.7-14.5] | 4.7-6.6 | 2.8-4.6 | < 2.8 | <0.001 |
| Tissue Doppler Lateral a' velocity, cm/s | 11.9<br>[6.8-17.0] | 4.3-6.7 | 1.7-4.2 | < 1.7 | 11.3<br>[6.4-16.3] | 3.9-6.3 | 1.4-3.8 | < 1.4 | 0.005 |
| Lateral E/e' ratio | 7.5<br>[2.7-12.3] | 12.4-14.6 | 14.7-17.0 | > 17.0 | 8.3<br>[3.8-12.7] | 12.8-14.9 | 15.0-17.2 | > 17.2 | <0.001 |
| Septal E/e' ratio | 9.1<br>[3.7-14.6] | 14.7-17.3 | 17.4-20.0 | > 20.0 | 9.6<br>[4.5-14.7] | 14.8-17.2 | 17.3-19.7 | > 19.7 | 0.033 |
| Average E/e' ratio | 8.3<br>[3.6-13.1] | 13.2 - 15.4 | 15.5-17.8 | > 17.8 | 8.9<br>[4.8-13.1] | 13.2-15.2 | 15.3-17.3 | > 17.3 | 0.001 |
| **Right Heart Structural and Functional Variables** |  |  |  |  |  |  |  |  |  |
| RA end systolic area, cm2 | 18.1<br>[10.6-25.6] | 25.7-29.3 | 29.4-33.1 | >33.1 | 14.6<br>[9.1-20.1] | 20.2-22.9 | 23.0-25.6 | >25.6 | <0.001 |
| RV end diastolic area, cm2 | 20.5<br>[13.2-27.8] | 27.9-31.4 | 31.5-35.1 | > 35.1 | 16.3 [10.9-21.6] | 21.7-24.2 | 24.3-26.9 | >26.9 | <0.001 |
| RV end diastolic area index, ml/m2 | 10.5<br>[7.3-13.7] | 13.8-15.3 | 15.4-16.9 | > 16.9 | 9.4<br>[6.7-12.1] | 12.2-13.5 | 13.6-14.8 | > 14.8 | <0.001 |
| RV end systolic area, cm2 | 12.5<br>[7.8-17.1] | 17.2-19.4 | 19.5-21.7 | > 21.7 | 9.5<br>[6.3-12.8] | 12.9-14.5 | 14.6-16.1 | >16.1 | <0.001 |
| RV end systolic area index, ml/m2 | 6.4<br>[4.3-8.5] | 8.6-9.6 | 9.7-10.6 | > 10.6 | 5.5<br>[3.9-7.1] | 7.2-7.9 | 8.0-8.7 | > 8.7 | <0.001 |
| Ratio of LV and RV end diastolic areas | 1.47<br>[1.0-1.9] | 0.8-0.9 | 0.6-0.7 | < 0.6 | 1.52<br>[1.1-1.9] | 0.9-1.0 | 0.7-0.8 | <0.7 | 0.006 |
| TAPSE, cm | 2.22<br>[1.54-2.89] | 1.20-1.53 | 0.87-1.19 | < 0.87 | 2.13<br>[1.49-2.76] | 1.17-1.48 | 0.85-1.16 | <0.85 | 0.001 |
| FAC, % | 39.2<br>[31.6-46.8] | 27.8-31.5 | 24.0-27.7 | < 24.0 | 41.3<br>[33.3-49.2] | 29.3-33.2 | 25.4-29.2 | <25.4 | <0.001 |
| Tissue Doppler S' Velocity, cm/s | 14.4<br>[8.8-20.0] | 6.0-8.7 | 3.2-5.9 | < 3.2 | 14.0<br>[8.2-19.8] | 5.3-8.1 | 2.4-5.2 | <2.4 | 0.072 |
| RV free wall e', cm/2 | 12.0<br>[6.0-18.0] | 3.1-5.9 | 0.1-3.0 | < 0.1 | 12.3<br>[5.6-19.0] | 2.2-5.5 | < 2.2* |  | 0.266 |
| RV free wall a', cm/s | 16.4<br>[8.5-24.4] | 4.5-8.4 | 0.6-4.4 | < 0.6 | 16.8<br>[9.1-24.5] | 5.2-9.0 | 1.4-5.1 | <1.4 | 0.319 |
| Tricuspid E-wave velocity, cm/s | 45.7<br>[25.0-66.5] | 66.6-76.9 | 77.0-87.2 | > 87.2 | 46.1<br>[28.7-63.5] | 63.6-72.1 | 72.2-80.8 | >80.8 | 0.634 |
| Tricuspid A-wave velocity, cm/s | 37.0<br>[19.9-54.0] | 54.1-62.6 | 62.7-71.1 | > 71.1 | 36.8<br>[19.4-54.2] | 54.3-62.9 | 63.0-71.6 | >71.6 | 0.796 |
| Tricuspid E wave deceleration time, | 194.9 | 298 - 348 | 349 - 399 | > 399 | 192.3 | 297-348 | 349-400 | >400 | 0.533 |
| msec | [93 – 297] |  |  |  | [88-296] |  |  |  |  |
| Tricuspid E/A ratio | 1.27<br>[0.7 - 1.8] | 1.9 - 2.1 | 2.2 - 2.4 | > 2.4 | 1.30<br>[0.7-1.9] | 2.0-2.2 | 2.3-2.5 | >2.5 | 0.181 |
| Tricuspid E/e' ratio | 4.0<br>[1.7 - 6.3] | 6.4 - 7.5 | 7.6 - 8.6 | > 8.6 | 4.0<br>[1.6-6.4] | 6.5-7.6 | 7.7-8.8 | >8.8 | 0.884 |
| Peak TR velocity, m/s | 2.6<br>[2.0-3.1] | 3.2-3.3 | 3.4 - 3.6 | > 3.6 | 2.5<br>[2.0-3.1] | 3.2-3.3 | 3.4 - 3.6 | >3.6 | 0.237 |
| RVSP, mmHg | 26.2<br>[15.2-37.1] | 37.2-42.6 | 42.7 - 48.1 | > 48.1 | 25.6<br>[14.5-36.8] | 36.9-42.4 | 42.5-48.0 | >48.0 | 0.256 |
| RA pressure, mmHg | 5.3 [2.7-7.9] | 8.0-9.2 | 9.3-10.5 | > 10.5 | 5.2<br>[3.0-7.4] | 7.5-8.5 | 8.6-9.6 | > 9.6 | 0.360 |
| PASP, mmHg | 31.5<br>[20.1-42.9] | 43.0-48.6 | 48.7 - 54.3 | > 54.3 | 30.9<br>[19.4-42.4] | 42.5-48.1 | 48.2-53.9 | >53.9 | 0.206 |
| IVC collapsibility, % | 65.7<br>[48.8-82.5] | 40.4-48.7 | 31.9-40.3 | < 31.9 | 66.8<br>[49.1-84.5] | 40.3-49.0 | 31.4-40.2 | < 31.4 | 0.124 |
| Maximum IVC diameter, cm | 1.52<br>[0.90-2.14] | 2.15-2.45 | 2.46 - 2.76 | > 2.76 | 1.46<br>[0.84-2.08] | 2.09-2.40 | 2.41-2.71 | >2.71 | 0.014 |
| Minimum IVC diameter, cm | 0.53<br>[0.17-0.89] | 0.90-1.07 | 1.08 - 1.25 | > 1.25 | 0.49<br>[0.12-0.86] | 0.87-1.04 | 1.05-1.23 | >1.23 | 0.011 |

**Table 3** reports differences in cardiac structure stratified by sex and race. In men, all four cardiac chambers were significantly larger in White participants and smallest in Chinese participants and persisted despite indexing for BSA. LV mass index, LV wall thickness, stroke work, IVC diameter, and tissue Doppler S’ velocity parameters were also significantly larger in White participants than in other racial/ethnic groups. Similar to men, LV volumes in women were smallest in Chinese and largest in White adults. However, unlike men, LVEF was highest in Chinese and lowest in White adults. Black women had the largest RV size, highest TAPSE, highest stroke work, and greatest LV wall thickness compared to other groups. In women, LV and RV diastolic parameters were similar amongst the different races/ethnicities. Hemodynamic measures did not differ substantially across races/ethnicities. **Figure 3** provides a graphical illustration of normative values stratified by sex and race/ethnicity in comparison to established thresholds.

**Figure 3.**
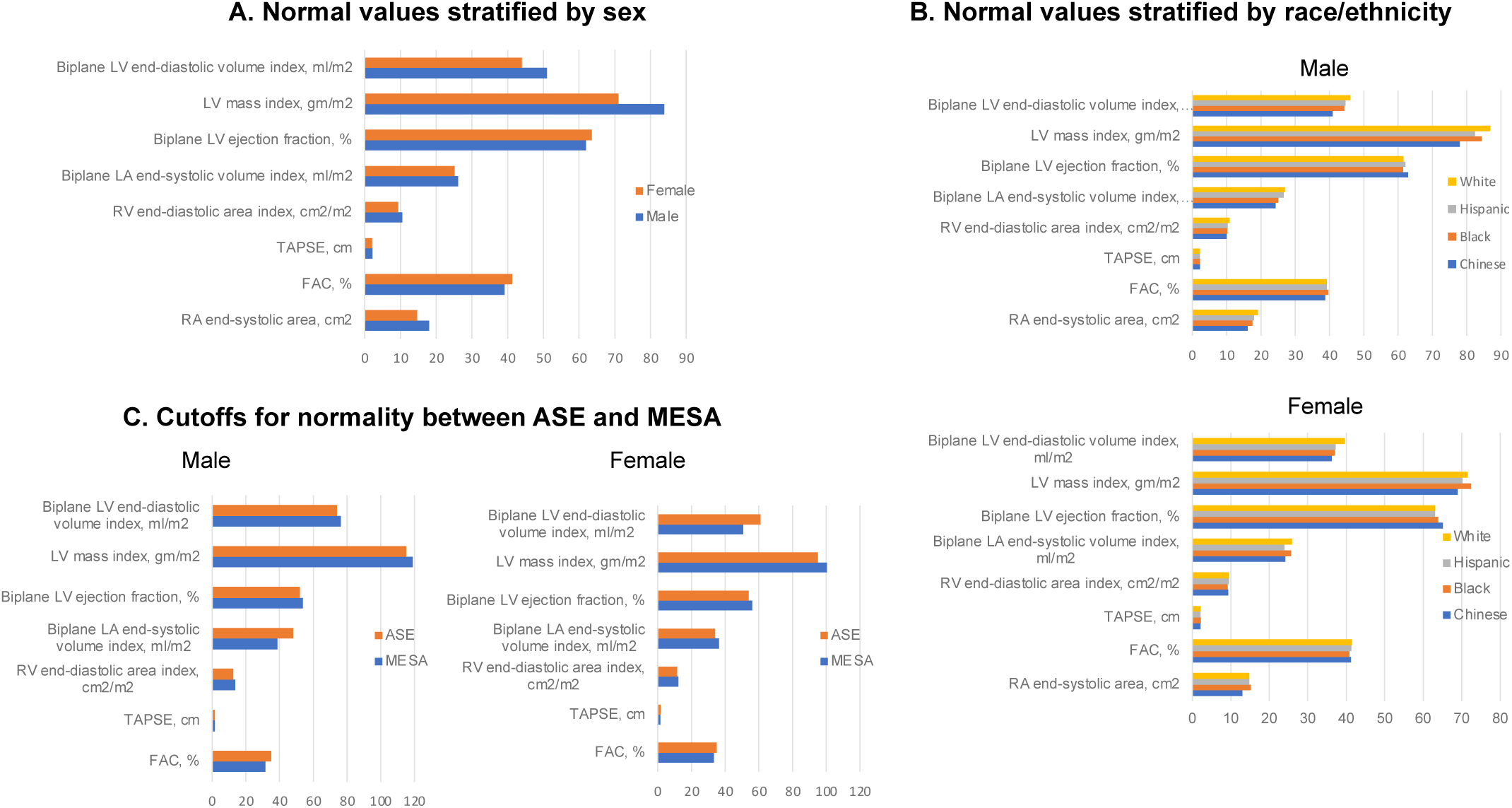
Outcome Measures by Included versus Non-Included Groups. Box and whisker plots show the distribution of functional outcome measures, including NT-proBNP in pg/mL. (**A**), 6-minute walk distance in meters (**B)**, and KCCQ overall summary score (**C)**. Lower NT-proBNP values and higher 6-minute walk distances and KCCQ scores indicate better functional status. Compared to the abnormal cohort (n=2424), those in the normal cohort (n=608) had lower NT-proBNP values, higher 6-minute walk distances, and higher KCCQ scores.

**Table 3.** Echocardiographic Characteristics of MESA Exam 6 Participants Stratified by Sex and Race/Ethnicity. Echocardiographic normal reference values MESA Exam 6 amongst the normal cohort (n = 608) stratified by race/ethnicity and self-reported sex. Values are listed as the mean [lower limit – upper limit of normal]. Lower and upper limits of normal were determined as the mean ± 2-3 standard deviations for all parameters. P-values for one-way analysis of variance tests for comparing mean parameter values across races/ethnicities are provided.

| Echocardiographic Parameters | MALES |  |  |  |  | FEMALES |  |  |  |  |
| --- | --- | --- | --- | --- | --- | --- | --- | --- | --- | --- |
|  | Chinese | Black | Hispanic | White | P-value | Chinese | Black | Hispanic | White | P-value |
| *Left Ventricular Structural and Functional Variables* |  |  |  |  |  |  |  |  |  |  |
| Biplane LA end-systolic volume, ml | 43.3<br>[20.7-65.9] | 50.1<br>[23.4-76.8] | 51.9<br>[27.6-76.1] | 54.0<br>[26.7 - 81.2] | <0.001 | 37.3<br>[21.9-52.7] | 47.9<br>[25.1 - 70.7] | 41.6<br>[24.8-58.4] | 45.8<br>[22.0-69.6] | <0.001 |
| Biplane LA end-systolic volume/BSA, ml/m2 | 24.3<br>[13.2-35.4] | 25.1<br>[11.2-39.0] | 26.7<br>[15.1-38.2] | 27.0<br>[14.1-39.9] | 0.04 | 24.2<br>[15.6-32.8] | 25.7<br>[13.8-37.7] | 24.0<br>[15.4-32.6] | 25.9 [14.0-37.7] | 0.07 |
| Biplane LV end-diastolic volume, ml | 72.7<br>[45.4-100.1] | 88.8<br>[51.0-126.5] | 86.9<br>[55.2-118.6] | 92.1<br>[57.8-126.4] | <0.001 | 55.8<br>[36.3-75.4] | 69.2<br>[42.4-96.0] | 64.5<br>[45.2-83.8] | 70.0<br>[43.5-96.5] | <0.001 |
| Biplane LV end-diastolic volume/BSA, ml/m2 | 40.9<br>[27.6-54.3] | 44.3<br>[27.3-61.3] | 44.7<br>[30.0-59.5] | 46.1<br>[30.6-61.6] | 0.001 | 36.3<br>[25.3-47.4] | 37.1<br>[23.4-50.8] | 37.3<br>[26.3-48.5] | 39.6<br>[27.4-51.8] | 0.002 |
| Biplane LV ejection fraction, % | 62.9<br>[55.7-70.1] | 61.4<br>[53.2-69.6] | 62.1<br>[53.7-70.5] | 61.6<br>[53.8-69.3] | 0.18 | 65.1<br>[57.3-73.0] | 63.9<br>[55.7-72.1] | 63.1<br>[56.6-69.5] | 63.1<br>[56.2-70.1] | 0.005 |
| Left ventricular end-diastolic dimension, cm | 4.2<br>[3.4-5.0] | 4.5<br>[3.5-5.5] | 4.4<br>[3.4-5.4] | 4.5<br>[3.5-5.5] | 0.044 | 3.9<br>[3.1-4.7] | 4.1<br>[3.1-5.1] | 4.2<br>[3.4-5.0] | 4.2<br>[3.4-5.0] | <0.001 |
| Left ventricular end-systolic dimension, cm | 2.8<br>[2.2-3.4] | 3.0<br>[2.2-3.8] | 3.0<br>[2.2-3.8] | 3.0<br>[2.2 - 3.8] | 0.19 | 2.5<br>[1.9-3.1] | 2.7<br>[1.9-3.5] | 2.8<br>[2.2-3.4] | 2.8<br>[2.2-3.4] | <0.001 |
| Left ventricular mass, gm | 139.0<br>[84.0-194.0] | 169.4<br>[86.2 - 252.6] | 160.4<br>[95.1-225.7] | 174.2<br>[90.5- 257.8] | <0.001 | 106.5<br>[60.9-152.0] | 135.1<br>[71.0-199.3] | 122.3<br>[65.4-179.2] | 126.8<br>[62.1-191.5] | <0.001 |
| Left ventricular mass index, gm/m2 | 78.0<br>[53.5-102.4] | 84.4<br>[46.4-122.4] | 82.5<br>[52.5-112.5] | 86.9<br>[49.1 -124.8] | 0.014 | 69.0<br>[44.7-93.3] | 72.4<br>[39.3-105.6] | 70.2<br>[43.7-96.7] | 71.5<br>[41.6-101.3] | 0.62 |
| Left ventricular outflow tract diameter, cm | 2.1<br>[1.9-2.4] | 2.2<br>[1.9-2.6] | 2.2<br>[1.9-2.5] | 2.3<br>[2.0 - 2.6] | <0.001 | 1.9<br>[1.6-2.1] | 2.0<br>[1.8-2.2] | 2.0<br>[1.7-2.3] | 2.0<br>[1.7-2.3] | <0.001 |
| Septal wall thickness, cm | 1.1<br>[0.8-1.5] | 1.2<br>[0.7-1.6] | 1.2<br>[0.8-1.5] | 1.2<br>[0.8-1.7] | 0.027 | 1.0<br>[0.7-1.3] | 1.1<br>[0.7-1.6] | 1.0<br>[0.6-1.4] | 1.0<br>[0.6-1.5] | 0.001 |
| Posterior wall thickness, cm | 0.9<br>[0.7-1.1] | 1.0<br>[0.7-1.2] | 1.0<br>[0.7-1.2] | 1.0<br>[0.71 - 1.2] | 0.003 | 0.8<br>[0.6 - 1.0] | 0.9<br>[0.7 - 1.1] | 0.8<br>[0.6 - 1.1] | 0.9<br>[0.6 - 1.1] | <0.001 |
| Fractional shortening, % | 33.3<br>[27.8-38.9] | 32.7<br>[24.1-41.2] | 32.2<br>[25.6-38.9] | 33.0<br>[25.4 - 40.6] | 0.38 | 35.2<br>[27.6-42.7] | 34.5<br>[26.9-42.0] | 33.6<br>[28.0-39.2] | 33.7<br>[26.8-40.0] | 0.007 |
| Stroke work, g*m | 76.4<br>[30.5-122.4] | 92.5 [39.5-145.4] | 87.6 [44.0-131.1] | 96.3 [50.5 - 142.1] | <0.001 | 60.0 [20.5-99.4] | 75.8 [36.0-115.5] | 68.2 [35.6-100.8] | 71.3 [33.1-109.5] | <0.001 |
| LVOT VTI, cm | 21.0<br>[12.4-29.6] | 21.7<br>[13.0-30.4] | 22.4 [14.4-30.3] | 22.6 [15.4 - 29.8] | 0.067 | 22.4 [13.5-31.4] | 24.0 [14.9-33.0] | 23.0 [14.6-31.4] | 23.2 [14.9-31.5] | 0.342 |
| Stroke volume, ml | 75.4<br>[37.6-113.3] | 84.4<br>[48.6-20.3] | 82.6<br>[49.9-115.3] | 90.0<br>[54.5-125.4] | <0.001 | 63.0<br>[34.2-91.7] | 75.1<br>[45.0-105.2] | 70.3<br>[40.5-100.1] | 72.1<br>[40.9-103.4] | <0.001 |
| Stroke volume index, ml/m2 | 42.3<br>[24.1-60.5] | 42.4<br>[23.0-61.7] | 42.6<br>[25.8-59.5] | 45.1<br>[27.9-62.4] | 0.11 | 41.3<br>[20.4-62.1] | 40.3<br>[24.9-55.7] | 40.6<br>[24.5-56.7] | 40.8<br>[25.7-55.9] | 0.94 |
| Cardiac output, L/min | 4.8<br>[2.2-7.5] | 5.2<br>[2.6-7.7] | 5.2<br>[2.7-7.7] | 5.4<br>[2.9-8.0] | 0.05 | 4.0<br>[2.0-6.1] | 4.6<br>[2.4-6.9] | 4.5<br>[2.5-6.5] | 4.6<br>[2.4-6.8] | 0.015 |
| Cardiac index, L/min/m2 | 2.7<br>[1.4-4.0] | 2.6<br>[1.4-3.8] | 2.7<br>[1.3-4.1] | 2.7<br>[1.6-3.9] | 0.77 | 2.7<br>[1.0-4.3] | 2.5<br>[1.3-3.6] | 2.6<br>[1.5-3.6] | 2.6<br>[1.5-3.8] | 0.45 |
| Left ventricular ejection time, ms | 310.0<br>[264.7-355.3] | 304.2<br>[257.3-351.1] | 307.4<br>[258.5-356.2] | 313.9<br>[266.3 - 361.4] | 0.12 | 314.7<br>[270.0-359.4] | 320.3<br>[273.1-367.4] | 319.1<br>[274.0-364.2] | 321.2 [279.7-362.7] | 0.34 |
| End-systolic elastance, mmHg/ml | 2.61<br>[1.35-3.87] | 2.40<br>[1.11-3.69] | 2.41<br>[1.34-3.47] | 2.19<br>[1.23-3.14] | <0.001 | 3.06<br>[1.87-4.25] | 2.69<br>[1.43-3.95] | 2.75<br>[1.18-4.31] | 2.74<br>[1.34-4.14] | 0.024 |
| Transmitral E-velocity, cm/s | 71.2<br>[39.6-102.9] | 71.4<br>[46.9-95.9] | 68.1<br>[39.4-96.7] | 68.4 [39.3-97.4] | 0.47 | 76.5 [43.7-109.4] | 81.6 [52.0-111.2] | 73.3 [43.2-103.4] | 75.8 [46.4-105.2] | 0.031 |
| Transmitral A-velocity, cm/s | 76.6 [35.2-117.9] | 76.6 [46.4-106.9] | 75.5 [41.7-109.2] | 73.1 [38.5-107.7] | 0.56 | 84.8 [50.9-118.6] | 82.3 [49.1-115.6] | 80.3 [41.3-119.3] | 78.8 [45.6-112.0] | 0.19 |
| E/A ratio | 0.97 [0.52-1.41] | 0.96 [0.50-1.42] | 0.94 [0.37-1.52] | 0.98 [0.41-1.54] | 0.89 | 0.93 [0.46-1.40] | 1.03 [0.44-1.63] | 0.97 [0.28-1.66] | 0.99 [0.52-1.46] | 0.28 |
| Mitral E-wave deceleration time, ms | 200 [126-274] | 201 [130-272] | 188 [112-264] | 211 [122-299] | 0.006 | 194 [115-273] | 190 [117-264] | 200 [116-283] | 192 [121-263] | 0.54 |
| Tissue Doppler Septal e' Velocity, cm/s | 8.0 [4.7-11.3] | 8.4 [4.4-12.5] | 7.3 [4.2-10.5] | 8.0 [4.2-11.9] | 0.028 | 8.1 [4.5 - 11.6] | 8.7 [4.6-12.7] | 7.8 [4.0-11.6] | 8.5 [4.3-12.8] | 0.037 |
| Tissue Doppler Lateral e' Velocity, cm/s | 9.5 [5.9-13.1] | 9.7 [5.3-14.1] | 9.7 [5.6-13.8] | 9.8 [4.7 - 15.0] | 0.88 | 9.1 [5.8 - 12.4] | 10.1 [5.3-14.8] | 9.7 [6.0-13.4] | 9.6 [5.3-13.9] | 0.15 |
| Tissue Doppler Septal a' Velocity, cm/s | 11.5 [8.4-14.6] | 11.5 [8.1-15.0] | 11.2 [8.0-14.4] | 11.5 [7.7 - 15.4] | 0.61 | 10.4 [7.4 - 13.4] | 10.7 [7.0-14.5] | 10.5 [6.5-14.6] | 10.7 [6.7-14.7] | 0.76 |
| Tissue Doppler Lateral a' Velocity, cm/s | 11.6 [6.9-16.3] | 10.9 [5.8-16.1] | 12.5 [7.1-17.1] | 12.0 [6.9 - 17.2] | 0.016 | 11.6 [6.7 - 16.6] | 11.3 [6.1-16.4] | 11.8 [7.2-16.3] | 11.1 [6.3-16.0] | 0.28 |
| Lateral E/e' ratio | 7.7 [3.4-12.0] | 7.7 [3.7-11.7] | 7.4 [2.7-12.0] | 7.4 [2.4-12.4] | 0.76 | 8.5 [5.1 - 12.0] | 8.4 [4.4-12.4] | 7.8 [3.9-11.7] | 8.3 [3.4-13.2] | 0.31 |
| Septal E/e' ratio | 9.2 [3.8-14.5] | 8.9 [4.2-13.5] | 9.6 [4.5 - 14.7] | 9.0 [3.4-14.6] | 0.45 | 9.7 [5.4-14.1] | 9.7 [5.8-13.6] | 9.9 [4.7-15.1] | 9.4 [4.0-14.8] | 0.60 |
| Average E/e' ratio | 8.4 [3.8-13.0] | 8.3 [4.5-12.1] | 8.5 [4.1- 12.9] | 8.2 [3.2-13.2] | 0.85 | 9.1 [5.7-12.5] | 9.0 [5.7-12.4] | 8.8 [4.8-12.9] | 8.9 [4.3-13.4] | 0.83 |
| **Right Heart Structural and Functional Variables** |  |  |  |  |  |  |  |  |  |  |
| RA end-systolic area, cm2 | 16.2 [9.7-22.7] | 17.5 [11.6-23.3] | 18.0 [10.5-25.5] | 19.1 [11.7-26.4] | <0.001 | 13.0 [8.8-17.3] | 15.2 [9.2-21.2] | 14.8 [10.2-19.5] | 14.8 [9.2- 20.3] | <0.001 |
| RV end diastolic area, cm2 | 17.8 [12.1-23.5] | 20.5 [13.8-27.2] | 20.0 [13.8-26.1] | 21.8 [14.8-28.8] | <0.001 | 14.2 [10.4-18.0] | 17.1 [12.0-22.7] | 16.3 [11.6-21.1] | 16.6 [11.5-21.7] | <0.001 |
| RV end diastolic area index, ml/m2 | 10.0 [7.2-12.9] | 10.2 [7.6- 12.8] | 10.3 [7.4-13.2] | 10.9 [7.6-14.3] | 0.001 | 9.3 [6.9-11.6] | 9.2 [6.5-11.9] | 9.5 [6.5-12.4] | 9.4 [6.9-12.0] | 0.59 |
| RV end systolic area, cm2 | 10.9 [7.0-14.8] | 12.4 [7.9-16.9] | 12.1 [8.2-16.1] | 13.3 [8.9-17.6] | <0.001 | 8.3 [6.1-10.5] | 10.1 [6.7- 13.6] | 9.6 [6.6-12.6] | 9.7 [6.6-12.8] | <0.001 |
| RV end systolic area index, ml/m2 | 6.2 [4.3-8.0] | 6.2 [4.4-8.0] | 6.2 [4.4-8.1] | 6.7 [4.5-8.8] | 0.003 | 5.4 [4.2- 6.7] | 5.4 [3.7-7.2] | 5.6 [3.8- 7.3] | 5.5 [3.9-7.1] | 0.81 |
| Ratio of LV and RV end diastolic areas | 1.5 [1.1-1.9] | 1.5 [1.1-1.9] | 1.5 [1.1 - 1.9] | 1.4 [1.0-1.9] | 0.041 | 1.6 [1.2-2.0] | 1.5 [1.0-2.0] | 1.5 [1.1-1.9] | 1.5 [1.1 - 1.9] | 0.41 |
| TAPSE, cm | 2.16 [1.51-2.80] | 2.21 [1.61 - 2.82] | 2.17 [1.53 - 2.82] | 2.27 [1.58 - 2.95] | 0.13 | 2.07 [1.51- 2.62] | 2.22 [1.59- 2.85] | 2.05 [1.53- 2.58] | 2.14 [1.48-2.81] | 0.016 |
| FAC, cm | 38.7 [31.5-46.0] | 39.7 [33.4- 46.0] | 39.2 [30.5- 48.0] | 39.2 [32.1- 46.4] | 0.70 | 41.2 [33.9- 48.5] | 40.8 [33.2- 48.4] | 41.3 [33.2- 49.4] | 41.4 [33.5-49.3] | 0.83 |
| Tissue Doppler S' Velocity, cm/s | 13.3 [8.5-18.1] | 14.3 [9.9-18.8] | 14.7 [8.4-20.9] | 14.8 [9.3-20.2] | 0.012 | 13.3 [8.8-17.9] | 14.0 [9.2- 18.9] | 13.4 [9.4-17.5] | 14.4 [7.7-21.0] | 0.054 |
| RV free wall e', cm/s | 11.4 [6.3 - 16.4] | 12.2 [7.5-17.0] | 11.7 [6.2-17.1] | 12.4 [5.9-18.9] | 0.13 | 11.7 [4.8-18.5] | 12.2 [7.1- 17.3] | 11.6 [6.8-16.5] | 12.8 [5.4-20.1] | 0.06 |
| RV free wall a', cm/s | 15.9 [8.1-23.6] | 17.0 [9.2- 24.7] | 16.0 [9.1-23.0] | 16.7 [8.6-24.9] | 0.38 | 16.4 [8.6-24.1] | 16.7 [8.4- 25.1] | 16.1 [9.9-22.2] | 17.2 [9.6-24.8] | 0.23 |
| Tricuspid E wave velocity, cm/s | 43.0 [27.1-59.0] | 44.6 [23.9- 65.2] | 45.8 [27.0- 64.7] | 47.1 [24.9- 69.2] | 0.10 | 44.8 [29.2- 60.4] | 45.6 [29.8- 61.4] | 48.0 [28.8- 67.2] | 45.9 [29.1-62.7] | 0.21 |
| Tricuspid A wave velocity, cm/s | 36.4 [21.7-51.2] | 36.1 [20.4- 51.8] | 38.9 [21.0- 56.9] | 36.6 [19.5- 53.6] | 0.27 | 36.8 [24.2- 49.4] | 35.9 [18.0- 53.7] | 39.5 [18.0- 61.1] | 36.0 [20.4-51.5] | 0.035 |
| Tricuspid E wave deceleration time, ms | 183 [91-276] | 207 [93-320] | 190 [94-286] | 199 [98-299] | 0.13 | 190 [103-277] | 213 [86-340] | 188 [95-281] | 188 [90-286] | 0.018 |
| Tricuspid E/A ratio | 1.2<br>[0.7-1.8] | 1.3<br>[0.8-1.7] | 1.2<br>[0.6-1.8] | 1.3 [0.8-1.9] | 0.063 | 1.3<br>[0.6-1.9] | 1.3<br>[0.6-2.0] | 1.3<br>[0.7-1.8] | 1.3<br>[0.8-1.9] | 0.36 |
| Tricuspid E/e' ratio | 4.0<br>[1.9-6.1] | 3.7<br>[1.9-5.5] | 4.1<br>[1.8-6.5] | 4.0 [1.7-6.4] | 0.43 | 4.1 [1.6 - 6.7] | 3.9<br>[1.6-6.2] | 4.3<br>[1.7-6.9] | 3.8<br>[1.6-6.1] | 0.042 |
| Peak TR velocity, m/s | 2.5<br>[2.0-3.1] | 2.6<br>[2.0-3.1] | 2.6<br>[2.1-3.0] | 2.5 [2.0-3.1] | 0.84 | 2.6<br>[2.1-3.1] | 2.5<br>[2.0-3.0] | 2.5<br>[2.0-3.1] | 2.5<br>[1.9-3.0] | 0.05 |
| RVSP, mmHg | 26.1 [14.5-37.8] | 26.5 [15.2-37.8] | 26.7 [17.3-36.1] | 25.9 [15.0-36.8] | 0.87 | 27.2 [16.9-37.5] | 25.9 [15.9-35.9] | 26.1 [14.8-37.5] | 24.8 [13.6-35.9] | 0.06 |
| RA pressure, mmHg | 5.2<br>[3.4-7.0] | 5.6<br>[2.4-8.8] | 5.1<br>[3.9-6.3] | 5.4 [2.4-8.4] | 0.21 | 5.0<br>[5.0-5.0] | 5.5<br>[2.5-8.5] | 5.2<br>[2.4-8.0] | 5.2<br>[3.2-7.2] | 0.18 |
| PASP, mmHg | 31.4 [19.5-43.2] | 32.1 [18.5-45.6] | 31.8 [22.5-41.1] | 31.4 [20.2-42.5] | 0.93 | 32.2 [21.9-42.5] | 31.4 [22.3-40.5] | 31.4 [19.0-43.9] | 30.0 [18.3 - 41.7] | 0.096 |
| IVC collapsibility, % | 65.6 [50.4-80.8] | 67.5 [52.2-82.8] | 65.1 [47.6-82.6] | 65.4 [48.3-82.4] | 0.57 | 67.6 [54.7-80.5] | 66.9 [49.7-84.1] | 64.7 [45.1-84.3] | 67.3 [49.6-84.9] | 0.24 |
| Maximum IVC diameter, cm | 1.41 [0.84-1.97] | 1.52 [0.76-2.28] | 1.46 [0.96-1.97] | 1.60 [1.02-2.18] | 0.001 | 1.30 [0.76-1.84] | 1.49 [0.84 - 2.14] | 1.42 [0.87 - 1.97] | 1.52 [0.91-2.12] | <0.001 |
| Minimum IVC diameter, cm | 0.49 [0.19-0.78] | 0.49 [0.17-0.82] | 0.51 [0.23-0.79] | 0.56 [0.16-0.97] | 0.02 | 0.42 [0.16-0.69] | 0.50 [0.13-0.87] | 0.51 [0.07-0.95] | 0.50 [0.15- 0.84] | 0.061 |

Notable differences in cutoffs for normative cardiac structure and function are observed compared to existing guidelines, as shown in **Table 4**. Overall, 496 (81.6%) individuals in the subgroup meeting our exclusion criteria would be classified as abnormal on at least one of these five parameters by existing ASE guidelines based on a younger population. Of the cohort meeting our exclusion criteria (n = 608), 36 (5.9%) individuals had a septal wall thickness ≥ 1.5 cm. Excluding these individuals, the mean septal wall thickness was 1.13 ± 0.17 cm in males and 1.00 ± 0.18 cm in females (p < 0.001), and the mean LV mass index was 82.3 ± 16.7 g/m^2^ in males and 70.3 ± 14.4 g/m^2^ in females (p < 0.001) and was not different across races/ethnicities, **Supplemental Table 3**. Using existing guidelines, 292 (51.0%) and 26 (4.5%) individuals would classify as having an abnormal septal wall thickness and LV mass index, respectively, and 447 (78.1%) would be classified as abnormal on at least one of the five parameters by existing ASE guidelines, **Supplemental Table 4**. **Table 5** demonstrates the association of selected echocardiographic parameters with the outcomes of all incident CVD, incident HF, and death utilizing ASE-vs. MESA-defined abnormal cut-off values based on AUC of Cox regression models. AUC p-values for testing between schemes were performed when p-values were significant. Findings demonstrate that AUC values for ASE-vs. MESA-defined abnormal cut-off values in the prediction of outcome are strongly dependent on the measure and outcome that are being evaluated. For example, several parameters such as LVEF and medial E/e’ have higher AUC when compared to MESA while AUC values were higher for peak TRV using ASE.

**Table 4.**
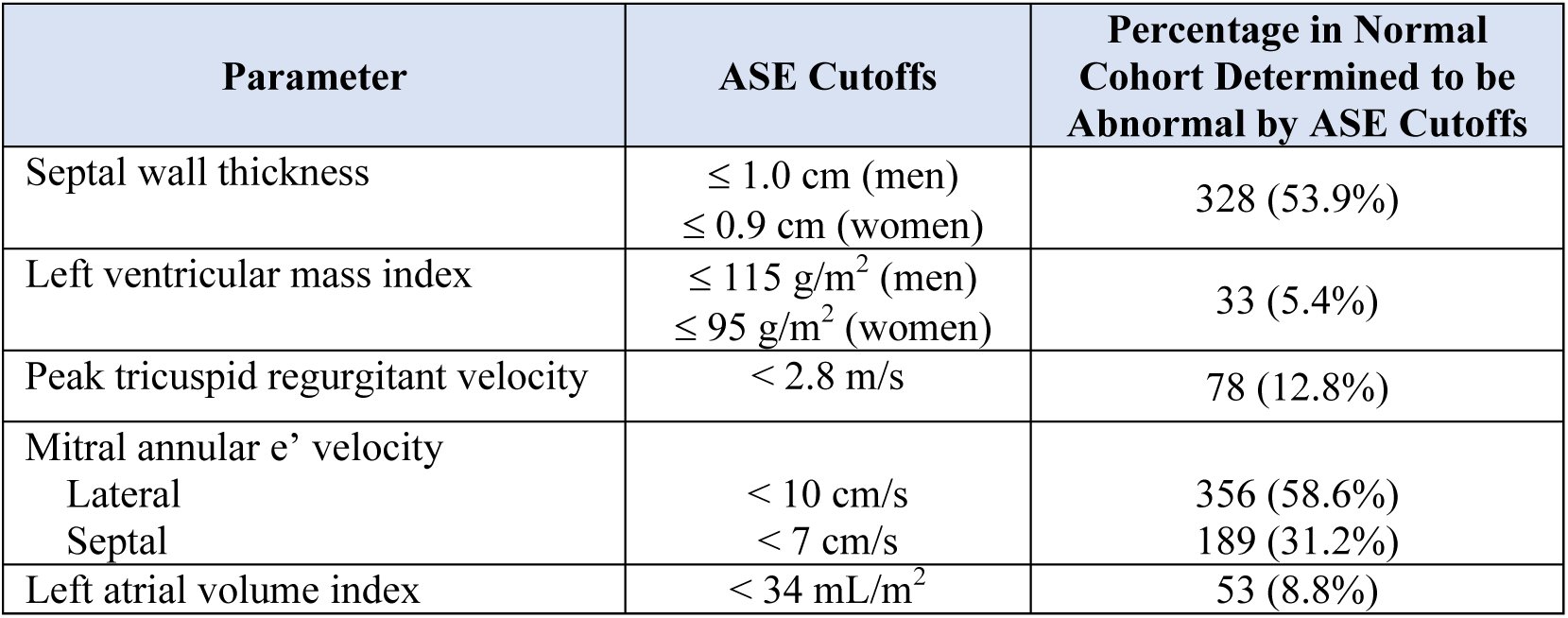
Percentage of Normative Cohort Classified as Abnormal based on Established Criteria. ASE Guideline Cutoffs for Normality is compared with five selected echocardiographic parameters measured as part of the MESA Exam 6 ancillary study and the number and percentage of patients in the normal cohort (n = 608) who would be determined to have abnormal values.

**Table 5.** Association of Selected Echocardiographic Parameters with Outcomes: Comparison of ASE-vs. MESA-defined abnormal cut-off values on Cox regression analysis. ASE and MESA AUC p-values: This is the p-value for testing if there is a significant difference in the Brier scores between the two models. The p-value from calculation of confusion matrix is Mcnemar’s Test p-values which test proportions of false positives and false negatives are equal.

| Echo Parameter | Outcome | ASE AUC | ASE AUC p-value | MESA AUC | MESA AUC p-value | ASE Sensitivity | ASE Specificity | ASE PPV | ASE NPV | ASE p-value | MESA Sensitivity | MESA Specificity | MESA PPV | MESA NPV | MESA p-value |
| --- | --- | --- | --- | --- | --- | --- | --- | --- | --- | --- | --- | --- | --- | --- | --- |
| LV end-diastolic volume index | All incident CVD* | 0.513 | 0.17 | 0.533 | 0.12 | 0.99 | 0.03 | 0.98 | 0.09 | <0.001 | 0.96 | 0.06 | 0.98 | 0.03 | <0.001 |
| LV end-diastolic volume index | Incident HF | 0.549 | 0.08 | 0.566 | 0.06 | 0.99 | 0.1 | 0.99 | 0.15 | 0.22 | 0.96 | 0.14 | 0.99 | 0.03 | <0.001 |
| LV end-diastolic volume index | Death | 0.511 | 0.09 | 0.553 | 0.003 | 0.99 | 0.03 | 0.96 | 0.11 | <0.001 | 0.96 | 0.11 | 0.97 | 0.09 | 0.039 |
| LV end-diastolic volume index | Incident HF or death | 0.52 | 0.021 | 0.557 | 0.006 | 0.99 | 0.05 | 0.96 | 0.21 | <0.001 | 0.96 | 0.12 | 0.96 | 0.11 | 0.44 |
| LV ejection fraction | All incident CVD* | 0.532 | 0.07 | 0.558 | 0.02 | 0.98 | 0.06 | 0.98 | 0.06 | 0.86 | 0.95 | 0.12 | 0.98 | 0.05 | <0.001 |
| LV ejection fraction | Incident HF | 0.602 | 0.014 | 0.663 | 0.003 | 0.98 | 0.21 | 0.99 | 0.08 | <0.001 | 0.95 | 0.31 | 0.99 | 0.05 | <0.001 |
| LV ejection fraction | Death | 0.559 | 0.001 | 0.573 | <0.001 | 0.97 | 0.13 | 0.97 | 0.15 | 0.29 | 0.94 | 0.16 | 0.97 | 0.1 | <0.001 |
| LV ejection fraction | Incident HF or death | 0.566 | <0.001 | 0.59 | <0.001 | 0.97 | 0.14 | 0.96 | 0.19 | 0.016 | 0.94 | 0.19 | 0.96 | 0.13 | 0.001 |
| Septal wall thickness | All incident CVD* | 0.478 | 0.58 | 0.6 | 0.06 | 0.16 | 0.95 | 0.99 | 0.03 | <0.001 | 0.91 | 0.14 | 0.98 | 0.04 | <0.001 |
| Septal wall thickness | Incident HF | 0.482 | 0.65 | 0.567 | 0.11 | 0.16 | 0.97 | 1 | 0.01 | <0.001 | 0.91 | 0.14 | 0.99 | 0.01 | <0.001 |
| Septal wall thickness | Death | 0.476 | 0.31 | 0.373 | 0.38 | 0.16 | 0.95 | 0.99 | 0.04 | <0.001 | 0.91 | 0.2 | 0.97 | 0.08 | <0.001 |
| Septal wall thickness | Incident HF or death | 0.475 | 0.51 | 0.599 | 0.011 | 0.16 | 0.95 | 0.99 | 0.05 | <0.001 | 0.91 | 0.18 | 0.96 | 0.08 | <0.001 |
| LV mass index | All incident CVD* | 0.681 | 0.005 | 0.635 | 0.019 | 0.86 | 0.3 | 0.98 | 0.05 | <0.001 | 0.9 | 0.21 | 0.98 | 0.05 | <0.001 |
| LV mass index | Incident HF | 0.796 | <0.001 | 0.729 | <0.001 | 0.87 | 0.59 | 1 | 0.04 | <0.001 | 0.9 | 0.45 | 0.99 | 0.04 | <0.001 |
| LV mass index | Death | 0.703 | <0.001 | 0.639 | <0.001 | 0.86 | 0.35 | 0.97 | 0.09 | <0.001 | 0.89 | 0.26 | 0.97 | 0.09 | <0.001 |
| LV mass index | Incident HF or death | 0.709 | <0.001 | 0.637 | 0.001 | 0.87 | 0.39 | 0.97 | 0.11 | <0.001 | 0.9 | 0.29 | 0.97 | 0.11 | <0.001 |
| Septal e' velocity | All incident CVD* | 0.856 | 0.004 | 0.591 | 0.001 | 0.56 | 0.69 | 0.99 | 0.04 | <0.001 | 0.96 | 0.2 | 0.98 | 0.11 | <0.001 |
| Septal e' velocity | Incident HF | 0.903 | 0.001 | 0.589 | 0.029 | 0.56 | 0.81 | 1 | 0.02 | <0.001 | 0.95 | 0.19 | 0.99 | 0.04 | <0.001 |
| Septal e' velocity | Death | 0.804 | <0.001 | 0.583 | 0.003 | 0.55 | 0.61 | 0.97 | 0.05 | <0.001 | 0.95 | 0.16 | 0.97 | 0.11 | 0.002 |
| Septal e' velocity | Incident HF or death | 0.83 | 0.009 | 0.587 | 0.004 | 0.55 | 0.65 | 0.97 | 0.06 | <0.001 | 0.95 | 0.16 | 0.96 | 0.13 | 0.06 |
| Lateral e' velocity | All incident CVD* | 0.908 | 0.041 | 0.602 | 0.05 | 0.3 | 0.8 | 0.98 | 0.03 | <0.001 | 0.95 | 0.14 | 0.98 | 0.07 | <0.001 |
| Lateral e' velocity | Incident HF | 0.983 | 0.005 | 0.551 | 0.11 | 0.29 | 0.96 | 1 | 0.01 | <0.001 | 0.95 | 0.11 | 0.99 | 0.02 | <0.001 |
| Lateral e' velocity | Death | 0.67 | 0.42 | 0.578 | 0.005 | 0.29 | 0.83 | 0.98 | 0.04 | <0.001 | 0.95 | 0.14 | 0.97 | 0.1 | 0.008 |
| Lateral e' velocity | Incident HF or death | 0.927 | 0.029 | 0.579 | 0.009 | 0.3 | 0.86 | 0.98 | 0.05 | <0.001 | 0.95 | 0.13 | 0.96 | 0.11 | 0.16 |
| E/e' ratio | All incident | 0.654 | <0.001 | 0.685 | <0.001 | 0.9 | 0.32 | 0.98 | 0.07 | <0.001 | 0.86 | 0.38 | 0.98 | 0.06 | <0.001 |
|  | CVD* |  |  |  |  |  |  |  |  |  |  |  |  |  |  |
| E/e' ratio | Incident HF | 0.701 | 0.001 | 0.773 | <0.001 | 0.9 | 0.41 | 0.99 | 0.04 | <0.001 | 0.86 | 0.56 | 1 | 0.04 | <0.001 |
| E/e' ratio | Death | 0.644 | <0.001 | 0.682 | <0.001 | 0.89 | 0.3 | 0.97 | 0.09 | <0.001 | 0.86 | 0.38 | 0.97 | 0.09 | <0.001 |
| E/e' ratio | Incident HF or death | 0.657 | <0.001 | 0.698 | <0.001 | 0.89 | 0.31 | 0.97 | 0.11 | <0.001 | 0.86 | 0.4 | 0.97 | 0.11 | <0.001 |
| LA volume index | All incident CVD* | 0.621 | 0.022 | 0.574 | 0.06 | 0.84 | 0.26 | 0.98 | 0.04 | <0.001 | 0.89 | 0.15 | 0.98 | 0.03 | <0.001 |
| LA volume index | Incident HF | 0.649 | 0.02 | 0.633 | 0.014 | 0.84 | 0.31 | 0.99 | 0.02 | <0.001 | 0.89 | 0.28 | 0.99 | 0.02 | <0.001 |
| LA volume index | Death | 0.669 | <0.001 | 0.612 | <0.001 | 0.83 | 0.37 | 0.97 | 0.08 | <0.001 | 0.88 | 0.26 | 0.97 | 0.08 | <0.001 |
| LA volume index | Incident HF or death | 0.67 | 0.001 | 0.617 | 0.003 | 0.83 | 0.35 | 0.97 | 0.09 | <0.001 | 0.88 | 0.26 | 0.96 | 0.09 | <0.001 |
| RV end-diastolic area index | All incident CVD* | 0.426 | 0.15 | 0.452 | 0.31 | 0.92 | 0.08 | 0.98 | 0.02 | <0.001 | 0.97 | 0.02 | 0.98 | 0.01 | 0.73 |
| RV end-diastolic area index | Incident HF | 0.613 | 0.024 | 0.517 | 0.35 | 0.92 | 0.21 | 0.99 | 0.02 | <0.001 | 0.98 | 0.03 | 0.99 | 0.01 | <0.001 |
| RV end-diastolic area index | Death | 0.564 | 0.004 | 0.525 | 0.017 | 0.92 | 0.15 | 0.97 | 0.06 | <0.001 | 0.97 | 0.06 | 0.97 | 0.09 | 0.035 |
| RV end-diastolic area index | Incident HF or death | 0.574 | 0.015 | 0.523 | 0.047 | 0.92 | 0.16 | 0.96 | 0.08 | <0.001 | 0.97 | 0.05 | 0.96 | 0.09 | 0.001 |
| TAPSE | All incident CVD* | 0.539 | 0.05 | 0.519 | 0.11 | 0.96 | 0.09 | 0.98 | 0.05 | <0.001 | 0.98 | 0.05 | 0.98 | 0.06 | 0.1 |
| TAPSE | Incident HF | 0.552 | 0.1 | 0.535 | 0.16 | 0.96 | 0.1 | 0.99 | 0.02 | <0.001 | 0.98 | 0.07 | 0.99 | 0.04 | 0.003 |
| TAPSE | Death | 0.556 | 0.002 | 0.542 | 0.002 | 0.95 | 0.13 | 0.97 | 0.09 | 0.002 | 0.98 | 0.1 | 0.97 | 0.15 | 0.003 |
| TAPSE | Incident HF or death | 0.553 | 0.01 | 0.542 | 0.004 | 0.95 | 0.12 | 0.96 | 0.09 | 0.08 | 0.98 | 0.09 | 0.96 | 0.17 | <0.001 |
| RV fractional area change | All incident CVD* | 0.474 | 0.34 | 0.52 | 0.16 | 0.91 | 0.06 | 0.98 | 0.02 | <0.001 | 0.97 | 0.05 | 0.98 | 0.04 | 0.21 |
| RV fractional area change | Incident HF | 0.597 | 0.044 | 0.535 | 0.17 | 0.91 | 0.17 | 0.99 | 0.02 | <0.001 | 0.97 | 0.07 | 0.99 | 0.02 | <0.001 |
| RV fractional area change | Death | 0.558 | 0.007 | 0.537 | 0.004 | 0.91 | 0.15 | 0.97 | 0.06 | <0.001 | 0.97 | 0.09 | 0.97 | 0.1 | 0.72 |
| RV fractional area change | Incident HF or death | 0.568 | 0.028 | 0.537 | 0.012 | 0.91 | 0.16 | 0.96 | 0.07 | <0.001 | 0.97 | 0.09 | 0.96 | 0.11 | 0.1 |
| Peak TR velocity | All incident CVD* | 0.645 | 0.03 | 0.566 | 0.014 | 0.75 | 0.32 | 0.98 | 0.03 | <0.001 | 0.96 | 0.14 | 0.98 | 0.07 | <0.001 |
| Peak TR velocity | Incident HF | 0.764 | 0.001 | 0.61 | 0.025 | 0.76 | 0.55 | 0.99 | 0.02 | <0.001 | 0.96 | 0.23 | 0.99 | 0.05 | <0.001 |
| Peak TR velocity | Death | 0.794 | <0.001 | 0.584 | 0.001 | 0.76 | 0.54 | 0.98 | 0.08 | <0.001 | 0.96 | 0.2 | 0.97 | 0.15 | 0.048 |
| Peak TR velocity | Incident HF or death | 0.781 | <0.001 | 0.588 | <0.001 | 0.76 | 0.53 | 0.97 | 0.09 | <0.001 | 0.96 | 0.2 | 0.96 | 0.18 | 0.46 |

## DISCUSSION

In a multi-ethnic representative cohort of older adults free of clinically overt CVD, we found numerous significant differences in cardiac structure and function across sexes and races/ethnicities. Moreover, we identified several clinically essential differences between current recommendations for normal limits for echocardiographic structure and functional variables and those encountered in a healthy older population. Specifically, we found that current guidelines may classify older individuals without cardiopulmonary or cardiometabolic disease as having abnormalities in cardiac structure and function in up to 81.6% of cases, despite these individuals having a better functional status than their peers and having no significant differences in risk for adverse outcomes.

Historically, recommended normative values ^24^ have resulted from at least two SD above and below the mean value derived from sizeable population-based cohort studies amongst young (18-45-year-old) individuals without prevalent CVD ^4, 32–34^. Comparatively few studies ^1–8^, have evaluated normative 2DE values for older adults with historical estimates predominantly derived from the Cardiovascular Health Study (CHS) ^1^. While the CHS study performed 2DE on 4,029 participants, the number of individuals ≥ 60 years for which normative 2DE data are available was limited to only 1,694 individuals. Further, while CHS demonstrated that several 2DE parameters vary with aging, few Hispanics and non-Black minorities were enrolled, the study utilized older non-digital media and reported several metrics no longer in clinical use.

More recently, the international World Alliance Societies of Echocardiography (WASE) study sought to establish 2DE reference ranges of cardiac chambers and function stratified by age, sex, and ethnicity ^2, 3^ employing standardized protocols for image acquisition with centralized expert interpretation at a single core site ^35^. However, including relatively younger adults, many Asian adults, raises the question of generalizability to an older, more diverse population. Additionally, while the WASE study excluded individuals with known CVD or those with greater than mild valvular dysfunction, adjudication was evaluated at a site level rather than using rigorous centrally determined inclusion criteria. Furthermore, prognostic information on those in the WASE cohort is lacking, precluding any information on whether parameters associate with improved outcomes or diminish the predictive value compared to the current ASE cutoff values.

In this large, multi-ethnic cohort of older adults without cardiopulmonary or cardiometabolic disease, after applying strict inclusion criteria, we found numerous differences in normative 2DE values by sex and race/ethnicity and clinically significant discrepancies with current chamber quantification guidelines. Notably, echocardiograms were performed by a dedicated core lab for research with multiple quality assurance checks applied, thus representing the gold standard for accurate, valid, and reproducible 2DE measures. We identified important differences in cardiac structure and function by sex. Specifically, we confirm that men have larger chamber sizes than women, despite accounting for differences in body habitus ^3^. By contrast, men had lower biventricular systolic function than women, with few exceptions. While biplane LVEF and fractional shortening were lower in men, stroke volume index and cardiac index were higher, indicating preload may be higher in men.

Similarly, despite no differences in TDI S’ and minimal differences in TAPSE across sexes, men had a lower RV FAC partly due to a larger RV end-diastolic area. These findings indicate the problems inherent in using ratio measures such as LVEF and FAC as markers of systolic function. Moreover, we confirm that, similar to the WASE study ^3^, women have smaller chamber dimensions and higher functional measures, even after accounting for body size, and this finding persists with age. In terms of diastolic function, while E/A did not differ across sexes, lower E-wave velocities as well as lower e’ velocities (particularly septal e’) in men resulted in higher septal and lateral E/e’ values in women. These changes suggest that females have less compliant ventricles than males with aging and may partially explain the female predominance of HF with preserved ejection fraction ^36^.

Similarly, we identified important differences in cardiac structure and function in older adults by race/ethnicity. Specifically, despite adjustment for body habitus, Chinese individuals had lower LV and LA volumes, LV mass, and RV and RA areas than non-Chinese individuals. By contrast, LV and RV functional measures were minimally different across races/ethnicities. Sex differences in measurements persisted despite stratification by race/ethnicity and independently confirmed those of the WASE study ^3^, suggesting that these differences persist in older adults. WASE Asian study participants were predominantly enrolled from international sites ^35^, and our results confirm that the differences observed are also seen in Asian Americans and, thus, not a function of site of enrollment or nationality. These findings further highlight the need for updated guidelines incorporating important chamber size and function differences by sex and race/ethnicity.

Additionally, we observed several significant discrepancies between established guidelines ^24^, predominantly based on a younger population and normative values in older adults. Importantly, we identified the lower limit of normal (defined as 1.96-SDs below the mean) for biplane LVEF to be 54.1% in men and 56.2% in women. Thus, it is plausible that so-called low-normal LVEFs (e.g., 50-55%) may be abnormal in an older population. Using these updated limits for normality, 15 (2.5%) of those in the normative aging cohort who lacked any cardiopulmonary or cardiometabolic disease would reclassify as having abnormal systolic function. Moreover, we find that a higher LV wall thickness, mass, LA volume index, and peak TR velocity may be expected in a healthy, older population and lower TDI mitral e’ velocities and not necessarily abnormal despite published guidelines ^24^.

A total of 81.6% of individuals in the normative aging cohort would be reclassified as having at least one abnormal finding using current guideline cutoffs for normality, including 53.9% septal wall thickness, 5.4% LV mass, 8.8% LA volume index, 12.8% peak TR velocity, and 63.3% TDI mitral e’ velocities. These changes may reflect subclinical changes in diastolic function with aging and thus raise the question of whether aging is truly physiologic. Importantly, the predictive capacity of ASE-versus MESA-derived cut-offs varied depending on the specific echocardiographic parameter and outcome under analysis. Taken together, these results indicate that age, sex, and race/ethnicity individually contribute significantly to risk prediction, and the predictive capacity fluctuates based on the particular parameter and outcome being assessed. Classifying these individuals as having abnormal findings based on normative values in a younger population may increase testing and anxiety due to applying a disease label. Thus, age should be strongly considered when classifying 2DE measurements as abnormal, and current guidelines should be updated to account for these critical age-related differences.

### Strengths and Limitations

The current study has several prominent strengths. First, our study uniquely represents participants across racial and ethnic groups. Second, referral bias is absent, as 2DE was performed for research purposes. Third, the influence of inter-rater and intra-rater variability is minimized due to comprehensive core lab adjudication of measurements. Fourth, as a cross-sectional study, temporal drift in measurements is absent. Fifth, including functional status information on participants, indicates that those individuals deemed normal differ from participants with abnormalities on outcomes of particular relevance to an aging population. Sixth, all outcomes were rigorously ascertained and independently adjudicated.

However, despite multiple strengths, there are several limitations to consider. First, MESA participants may be healthier than non-participants due to volunteer bias, and upon entry into the MESA study, the absence of clinically overt CVD was an exclusion criterion. Furthermore, there may be survival bias with healthier participants remaining in the study until exam 6, approximately 16 years after the baseline exam. However, these characteristics of the MESA participants may represent strength in identifying a healthy aged population, despite a minority of patients ≥ 85 years. Second, while strict, comprehensive inclusion criteria defined the normal cohort, it is still possible that unmeasured subclinical abnormalities may exist in the subgroup meeting our exclusion criteria. Third, as MESA enrolled an older cohort, these results should not be applied to a younger population. Fourth, septal wall thickness up to 1.6 cm (men) and 1.5 cm (women) was noted and may reflect the frequency to which basal septal wall thickening is encountered in older adults. However, despite excluding individuals with a septal wall thickness ≥1.5 cm, for whom it is impossible to exclude hypertrophic cardiomyopathy without further testing ^37^, overall results were substantially unchanged. Lastly, the impact of indexation on parameters such as LV mass ^38^ and classification of LVH severity ^39^ derived from cardiac magnetic resonance imaging has previously been shown to be limited in the MESA cohort, given the diversity of body size in these participants. Future studies designed to evaluate the impact of indexation on echocardiographic parameters are an area of active investigation.

## Conclusions

In a multi-ethnic, population-based cohort of older adults free of known, clinically overt cardiopulmonary or cardiometabolic disease, we found numerous significant differences in cardiac structure and function across sexes and races/ethnicities. Based on younger individuals, current guidelines classify up to 81.6% of healthy older adults as abnormal. Our findings suggest that current guidelines for determining abnormality should be revised to account for sex-specific differences by race/ethnicity that occur with healthy aging.

### ABBREVIATIONS, *alphabetical order*

2DE: Two-Dimensional Echocardiography
6MWD: Six-Minute Walk Distance
ASE: American Society of Echocardiography
AUC: Area Under the Curve
AVA: Aortic Valve Area
BMI: Body Mass Index
BPM: Beats Per Minute
CHS: Cardiovascular Health Study
CKD: Chronic Kidney Disease
COL: Columbia University, New York, NY
COPD: Chronic Obstructive Pulmonary Disease
CVD: Cardiovascular Disease
eGRF: Estimated Glomerular Filtration Rate
FAC: Fractional Area Change
HF: Heart Failure
HTN: Hypertension
ICC: Intraclass Correlation Coefficients
IVC: Inferior Vena Cava
JHU: Johns Hopkins University, Baltimore, MD
KCCQ: Kansas City Cardiomyopathy Questionnaire
LV: Left Ventricle
LVEF: Left Ventricle Ejection Fraction
LVOT: Left Ventricular Outflow Track
MESA: Multi-Ethnic Study of Atherosclerosis
NPV: Negative Predictive Value
NT-proBNP: N-terminal pro-B-type Natriuretic Peptide
NWU: Northwestern University, Chicago, IL
PASP: Pulmonary Artery Systolic Pressure
PPV: Positive Predictive Value
RA: Right Atrium
RV: Right Ventricle
RVSP: Right Ventricular Systolic Pressure
SBP: Systolic Blood Pressure
SD: Standard Deviation
TAPSE: Tricuspid Annular Plane Systolic Excursion
TDI: Tissue Doppler Imaging
TR: Tricuspid Regurgitation
UCLA: University of California, Los Angeles, CA
UMN: University of Minnesota, Minneapolis, MN
VTI: Velocity Time Integral
WASE: World Alliance Societies of Echocardiography
WFU: Wake Forest University, Winston-Salem, NC

## Data Availability

All MESA data is available by request through www.mesa-nhlbi.org.

## Acknowledgments.

The authors thank the other investigators, the staff, and the MESA participants for their valuable contributions. A complete list of participating MESA investigators and institutions is available at http://www.mesa-nhlbi.org.

## Author Contributions

SJS and AB were principal investigators of the MESA Early Heart Failure Study (which funded the echocardiograms, 6MWD, and KCCQ at MESA Exam 6), and SJS and LN designed the MESA echocardiographic protocol. MM, JA, JBS, and BF designed the analytic plan for this study. MH and JBS performed the statistical analysis. MM and JBS wrote the initial draft. All other authors provided critical revisions for important intellectual content. All authors approved the final draft for submission.

## Clinical Perspectives

- Healthy aging is associated with sex-specific changes to cardiac chamber size and function.
- Changes in cardiac chamber size and function can be seen in healthy older cardiopulmonary and/or cardiometabolic disease, stratified by sex and racial/ethnic groups.
- Current echocardiographic guidelines may incorrectly classify healthy older adults as having abnormal parameters.
- Future echocardiographic guidelines and reporting standards should consider normal healthy aging, age, sex, and racial/ethnic groups.

## Central Illustration

Within a well-characterized cohort of older adults, we identified 608 patients without cardiopulmonary disease and found numerous differences in cardiac chamber size and function, when stratified by sex and race/ethnicity. Current guidelines should adjust for normal physiologic aging.

---

**Supplemental Table 1.** Cutoff Values Used for Defining Abnormality

**Supplemental Table 2.** Results of multivariable regression models.

**Supplemental Table 3.** Sensitivity analysis results excluding individuals with a septal wall thickness ≥ 1.5 cm on normal reference values for septal wall thickness and mass stratified by race/ethnicity and sex.

**Supplemental Table 4.** Results of sensitivity analysis excluding individuals with a septal wall thickness ≥ 1.5 cm compared with ASE guideline cutoffs for normality.

